# Quicker team launch times for urgent priority neonatal retrievals: A Quality Improvement Initiative study

**DOI:** 10.1101/2024.10.28.24316327

**Authors:** Saumil A Desai, Kevin George, Kylie McDonald, Alysha Timoney, Dahna Kelland, Stephanie Barr, Molly Carroll, David Lockhart, Olivia Peters, Jonathan Davis

## Abstract

**Background:** Neonatal retrieval networks developed time-centric quality metrics as Key Performance Indicators (KPI) for benchmarking recently. Quicker launch time (departure from base), an essential KPI, enables neonatal retrieval teams to rapidly provide higher-level care to sick infants. Newborn Emergency Transport Services, Western Australia (NETS WA) facilitates neonatal transfers across largest global retrieval area necessitating quicker team launch times for urgent retrievals. NETS WA conducted Quality Improvement (QI) study to quicken team launch times for urgent priority retrievals.

**Aims:** Smart aim was to quicken NETS WA team launch times on urgent priority retrievals to ≤15 minutes (Australian New Zealand Neonatal Retrieval Network 2022). Secondary aims included impact of quicker launch times on time-centric quality metrics (“first look time”, “total retrieval time”).

**Settings:** This study was completed over two years by NETS WA. Urgent priority retrievals comprise 10-15% of total transfers (∼1200/year).

**Interventions:** QI team introduced Plan-Do-Study-Action (PDSA) cycles: 1. immediate access to transport cots 2. additional personnel 3. pre-defined priority matrix 4. direct communication strategies. Quality metrics (launch time, first look time, total retrieval time) were gathered from electronic retrieval database (REDCap). Data collection done at baseline (January-May 2022), during PDSA cycles (June 2022-April 2023) and post last PDSA cycle (May-December 2023).

**Results:** Launch times reduced from median (IQR) duration of 35.5 (21.5-90.0) to 17.0 (11.0-37.0) minutes for all urgent priority retrievals (p 0.0006). Launch times for road only urgent retrievals reduced to the recommended median (IQR) duration of 15.0 (10.0-20.0) minutes (p 0.009). Quicker launch times reduced first look time from median (IQR) of 85.0 (54.8-269.3) to 52.5 (30.5-152.3) minutes (p 0.008). Total retrieval time reduced from median (IQR) of 243.5 (135.8-395.3) to 182.0 (117.0-390.0) minutes (p 0.33).

**Conclusion:** Well-designed QI measures enabled NETS WA teams to quicken essential time-centric quality metrics for urgent priority neonatal retrievals.

**What is already known on this topic:**

- Quicker launch times are crucial for retrieval teams for rapid access provision of higher-level care to sick neonatal infants.
- There is dearth of literature on measures to reduce launch times of neonatal retrieval teams.

**What this study adds:**

- Newborn Emergency Transport Services Western Australia (NETS WA) Quality Improvement (QI) measures quickened team launch times on urgent priority neonatal retrievals.
- Quicker launch times improved other essential time-centric quality metrics (“first look time” and “total retrieval time”) for urgent priority neonatal retrievals.

**How this study might affect research, practice or policy:**

- The study provides data for consensus-based time-centric quality metrics for national and international benchmarking.
- Generalizable study QI measures for other neonatal retrieval teams to adopt and reduce their launch times.

## Introduction

Centralization of health care require neonatal retrieval teams to provide higher level care to sick or premature infants in distant settings^1^. Approximately, 8-25% of neonatal retrievals with an imminent threat to life or organ are time-critical^2, 3^. These retrievals necessitate the specialist neonatal teams to attend as rapidly as possible. Delays in the retrieval team departure from base can consequently impact the provision of this time-critical care ^4^.

The retrieval team activation is variably referred to as “mobilization time” by Ground and Air Medical Quality Transport (GAMUT 2015) database and “launch time” by the Australian New Zealand Neonatal Retrieval Network (ANZNRN 2022) data dictionary. This is an important time-centric quality metric indicator for benchmarking of retrieval teams ^5–7^. Quicker launch times (“time taken from dispatch to leave base”) prove more beneficial than minimizing stabilization times (“time from first look at baby till ready to leave”) for urgent priority neonatal retrievals^4, 7, 8^. This is due to unique neonatal physiology requiring longer stabilization compared to adult population^9^. Launch times are influenced by several factors such as availability of a trained retrieval team 24x7, in-house ambulance service, triaging, functional equipment ready for deployment and communication education ^4, 10^.

The Newborn Emergency Transport Services, Western Australia (NETS WA) is the sole retrieval service for transferring neonatal infants across the state (2.6 million sq kms), the largest retrieval area in the world operated by a single team ^11–13^. NETS WA has prolonged retrieval durations and quicker launch times are crucial to attend to sick infants in a time-critical manner. The present study aims to introduce QI initiatives to quicken the NETS WA team launch times on urgent priority retrievals.

## Methods

### Study setting

WA has approximately 34,000 live births annually^11^; 3-4 % (1100-1200 per year)^12, 13^ of these infants are transferred by NETS WA to one of two tertiary neonatal intensive care units (NICU) in the state capital, Perth. The furthest trips are around 2200km and may take ∼24 hours to complete^12–14^. Each NETS WA retrieval team comprises of doctors (senior registrar and/or neonatal consultant), nurses (neonatal trained) and paramedical staff (Ambulance Transport Officer). NETS WA team operates with support from St John’s Ambulance services for Perth metropolitan urban area that sprawls over 175 kms (road only). Approximately ∼70% of the NETS WA retrievals are within the Perth metropolitan area. NETS WA is supported by the Royal Flying Doctor Services Western Operations (RFDS) for transfers outside the metropolitan area (∼30%) that require rotary and fixed wing aircrafts for retrievals (road and air)^12–14^. The usual process of NETS WA retrievals is like other standard neonatal retrieval teams (appendix 1). The Child and Adolescent Health services (CAHS) neonatal directorate encompasses the tertiary perinatal and surgical neonatal units at King Edward Memorial Hospital (KEMH) and Perth Children’s Hospital (PCH) respectively, apart from NETS WA.

### Ethics approval

This study was conducted after approval from the institutional committee for QI projects (Governance Evidence Knowledge Outcomes, GEKO WA project number 50894). A waiver of ethics was granted. The study followed the standard SQUIRE 2.0 guidelines ^15^.

### Design

This was a single centre QI study undertaken by NETS WA team.

### Definitions

Launch time is the “Time the team departed base” by the ANZNRN data dictionary (2022)^7^. The dictionary defines the “urgent” retrievals as those that should have launch times of less than 15 minutes. The “serious” retrievals are classified as those requiring launch times between 30-60 minutes and those classified as “Not time critical” could be more than 60 minutes^7^. The other time-centric quality metrics evaluated were “first look time” and “total retrieval time”. “First look time” is defined as “time the NETS team first saw the patient” and the “total retrieval time” is the “total time taken from the initial referral call till the retrieval was completed” (ANZNRN 2022) ^7^.

### Aims and goals

The primary aim was to quicken the launch times of the NETS WA teams for urgent transfers. The SMART (Specific, Measurable, Achievable, Relevant, and Time-Bound) goal of this QI study was to reduce the launch times of the NETS WA teams for urgent retrievals to 15 minutes or less (recommended by ANZNRN 2022) ^7^. The secondary aim was to evaluate the impact of quicker team launch times on the “first look time” and “total retrieval time”.

#### Study period

The study was conducted over two years (January 2022-December 2023).

#### Data collection

The study variables are described in appendix 2. These variables are routinely entered by NETS WA team members onto the Research Electronic Data Capture (REDCap) web application during each retrieval. Data was collected from before, during and after the PDSA cycles in the study period.

### Quality Improvement (QI) team

A core NETS WA QI team comprising of the medical director, retrieval consultant, neonatal senior registrars, clinical nurse consultant and senior neonatal nurses was formed.

### Baseline period (January-May 2022)

Focussed group discussions (FGD) were conducted to review local retrospective audits and existing literature at baseline. The data collected in this period determined the time-centric metrics of NETS WA teams on urgent priority retrievals at baseline. The group conducted an online poll with the team members to understand the factors impacting the current launch times. This delineated the root cause analysis and modifiable risk factors in a driver diagram (figure 1). Consensus-based QI initiatives were introduced as the Plan-Do-Study-Action (PDSA) cycles in timely intervals to address these factors. Plan-Do-Study-Action cycles as central component of QI initiatives have previously proven impactful in neonatal care^16, 17^. Four PDSA cycles as part of QI initiatives were introduced during the study period to achieve the SMART goal. FGD were organized at the beginning of each PDSA cycle before implementation. FGD were organized after the last PDSA cycle to ensure the impact was sustained in the long run.

**Figure 1:**
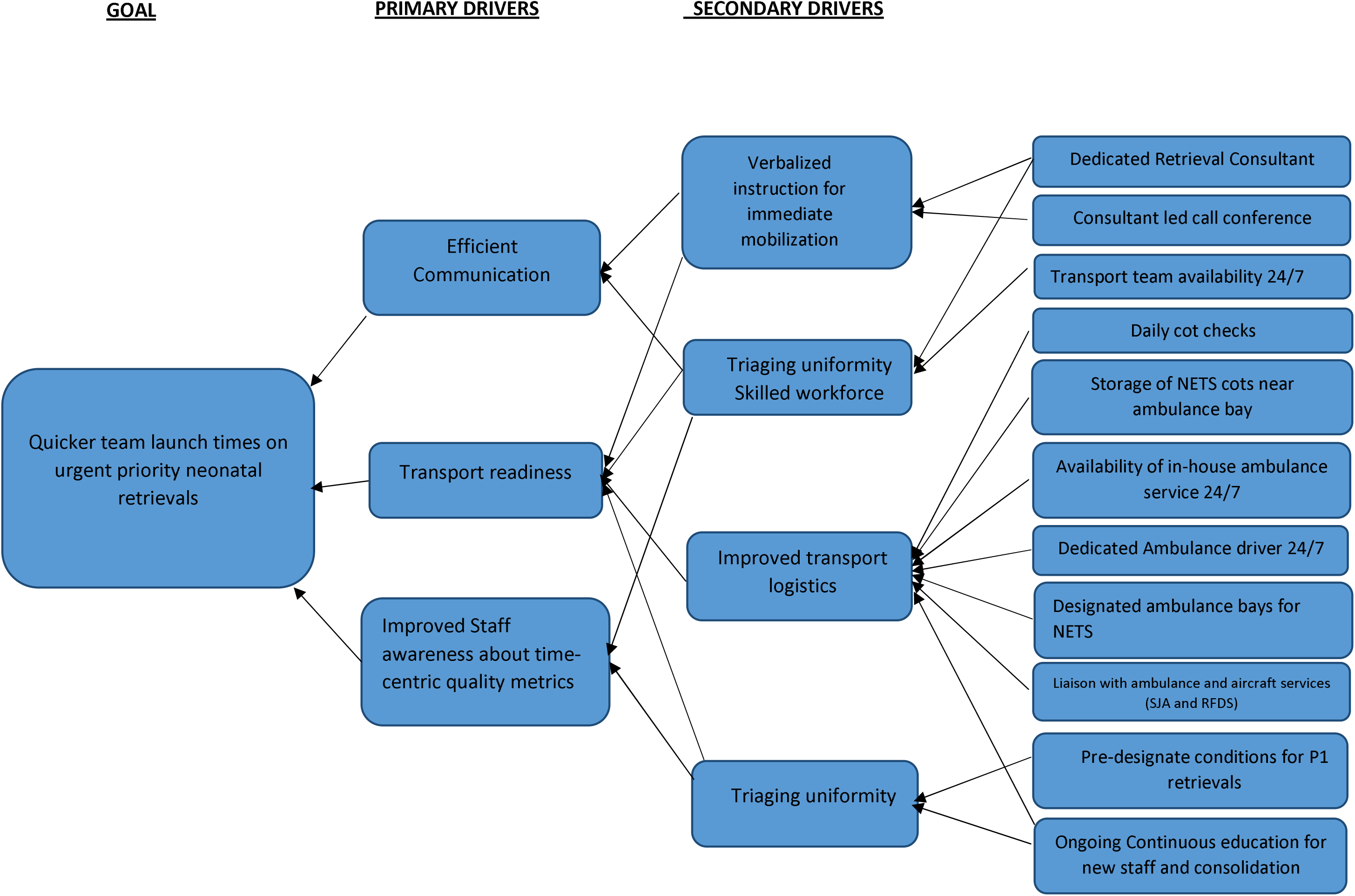
Driver diagram

The different PDSA cycles with their timeline are detailed below:

### PDSA cycle 1 (June 2022)

#### Equipment

Storage of NETS WA cots in a secure room next to the ambulance bays. Previously, the NETS WA cots were stored in the NETS WA office located on level 3 in PCH. The mobilization of NETS WA cots from level 3 to the ambulance for loading would approximately take 3-4 minutes. This duration was targeted for shortening by storing the cots in a secure room next to the ambulance bays in the first PDSA cycle.

#### Education

Weekly sessions to educate staff about NETS specific processes. The junior medical and nursing staff rotate every few months between the different CAHS neonatal directorate sites (NETS WA, KEMH and PCH) mentioned previously. Hence, regular education sessions were conducted to orientate the new rotating staff about the definitions and expected durations of time-centric quality metrics as per the ANZNRN data dictionary^7^ appendix 2)

### PDSA cycle 2 (October 2022)

Personnel: NETS WA referral calls were previously (until October 2022) led by the PCH NICU consultants for management, in addition to their existing ward responsibilities. The newly recruited NETS WA only consultants were responsible for leading retrieval calls, management and accompanying retrieval teams without any ward responsibilities. The QI team consolidated the NETS processes and education for time-centric quality metrics with the help of these dedicated neonatal retrieval consultants^7^.

### PDSA cycle 3 (February 2023)

#### Triage uniformity

A set of conditions (medical and surgical) were designated as automatic activation for P1 retrieval tasking of NETS WA teams from February 2023 (appendix 3). This led to uniformity in tasking for P1 retrievals. Before this time, P1 tasking was done at the discretion of the retrieval consultant.

#### Ongoing Education

Weekly sessions to educate new staff about NETS specific processes and time-centric quality metrics, as elaborated in PDSA cycle 1 (appendix 2).

### PDSA cycle 4 (April 2023)

#### Verbal Communication effectiveness

Communicate clearly on the NETS WA call conference system to designate the selected condition as P1. To instruct the team to actively leave the call-conference and get ready to depart the NETS WA base within 15 minutes of this instruction.

#### Ongoing Education

Weekly sessions to educate new rotating staff about NETS specific processes and time-centric quality metrics, as elaborated in PDSA cycle 1 (appendix 2).

#### Post-PDSA period (May-December 2023)

Data was collected for another 8 months to ensure that the improvement was sustained beyond the last PDSA cycle.

The study flow chart with timeline for PDSA cycles and data collection is depicted in figure 2.

**Figure 2:**
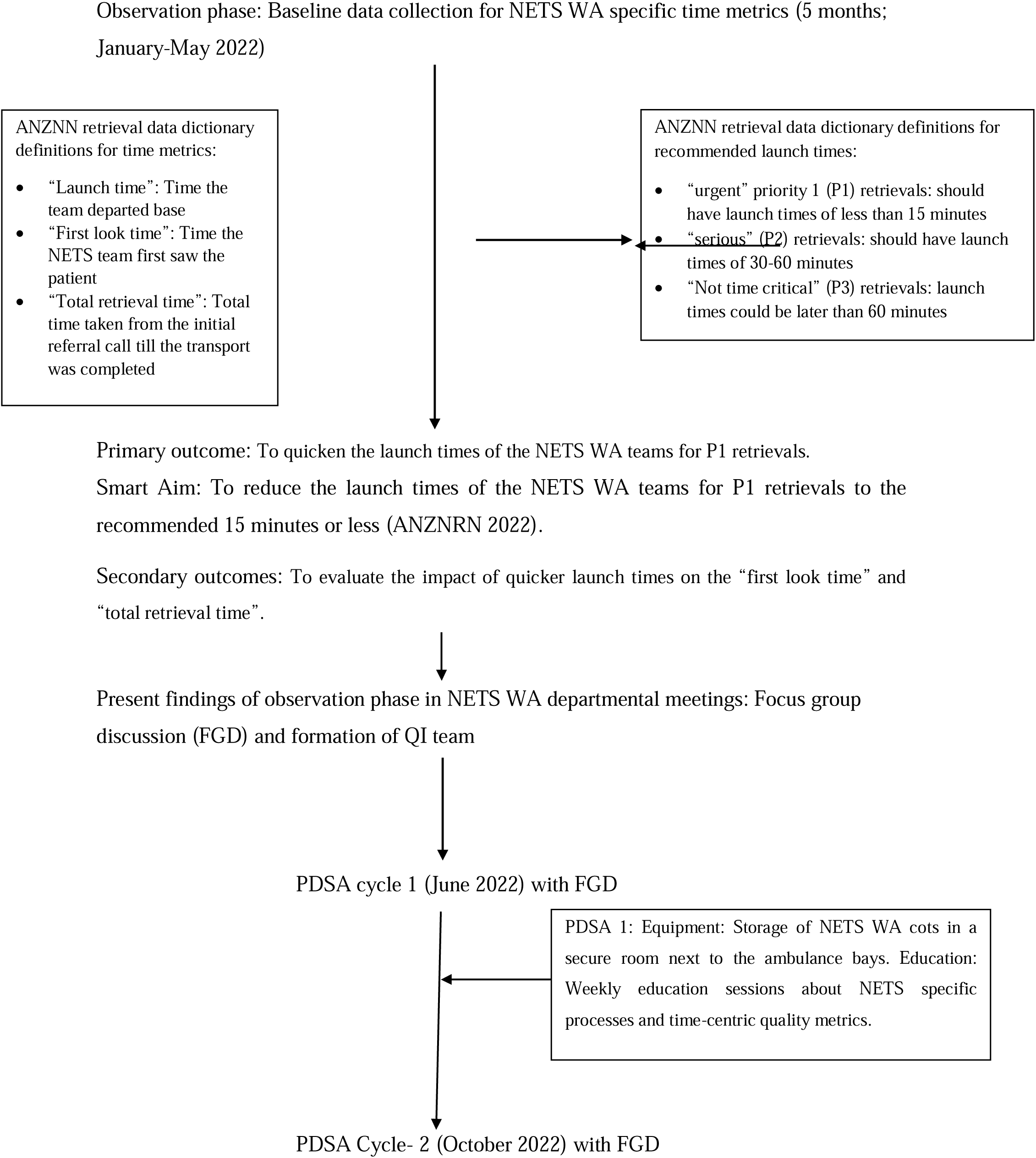
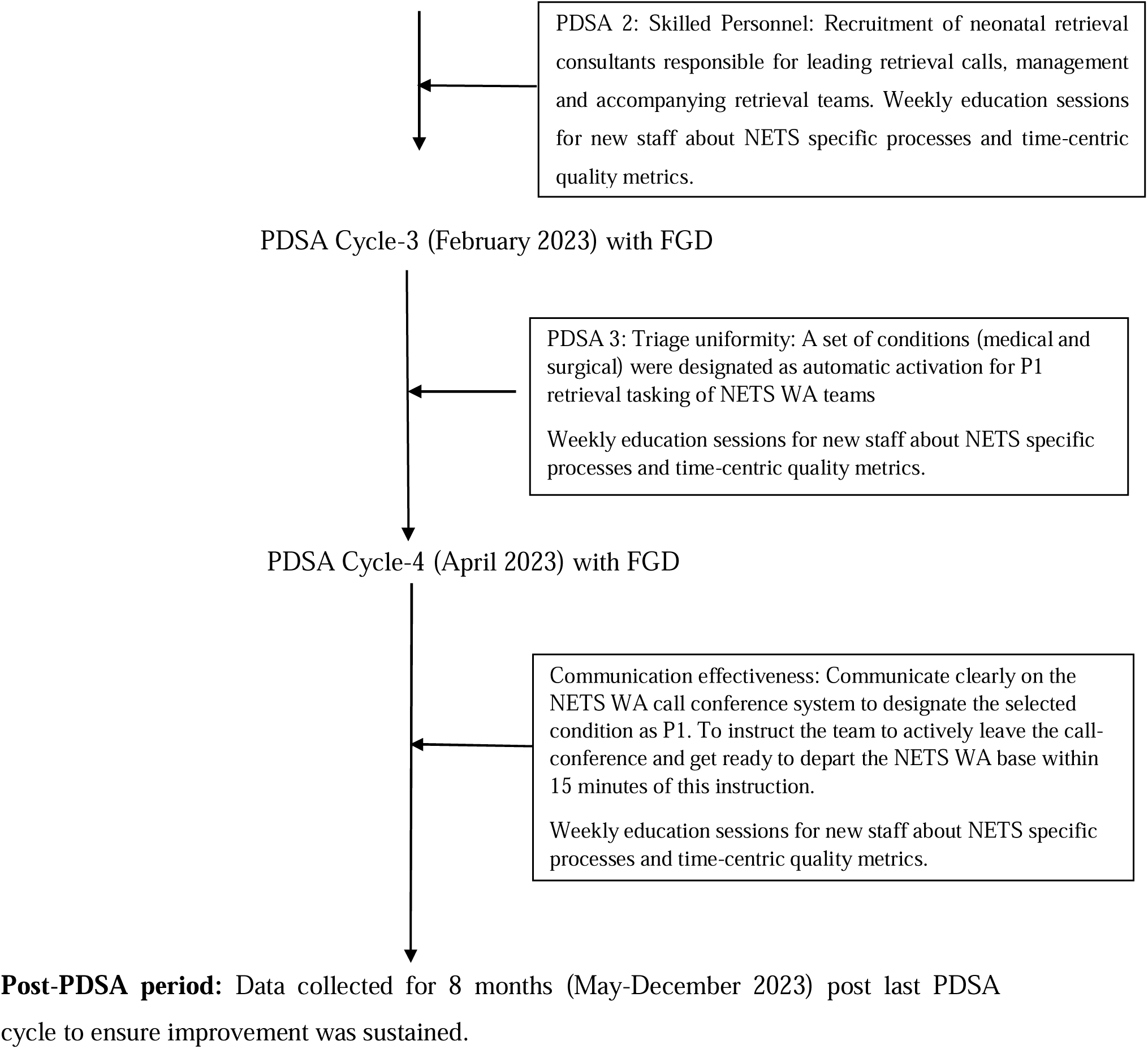
Study flow diagram

### Statistical analysis

The data variables for time-centric metrics were compared for three epochs ie baseline: Jan-May 2022, PDSA period: June 2022-April 2023, post PDSA period May-December 2023.

Categorical variables were summarized using frequency and percentage and compared using the Fisher’s exact test. Continuous variables were expressed as median and inter-quartile ranges (IQR) if non-parametric. Groups were compared using Kruskal-Wallis test with Dunn’s correction for multiple comparisons between the 3 epochs. A p value of <0.05 was considered statistically significant. Statistical analysis was performed using the GraphPad Prism (MA, USA).

The study period of 24 months (January 2022-December 2023) was divided into 2-month blocks for the interrupted time series analysis. Analysis was extended for 8 months post the last PDSA cycle. This additional analysis was important to ascertain whether the adherence to time-centric metrics was sustained. Run charts were created to describe median durations of the time-centric quality metrics over the study period (figure 3).

**Figure 3:**
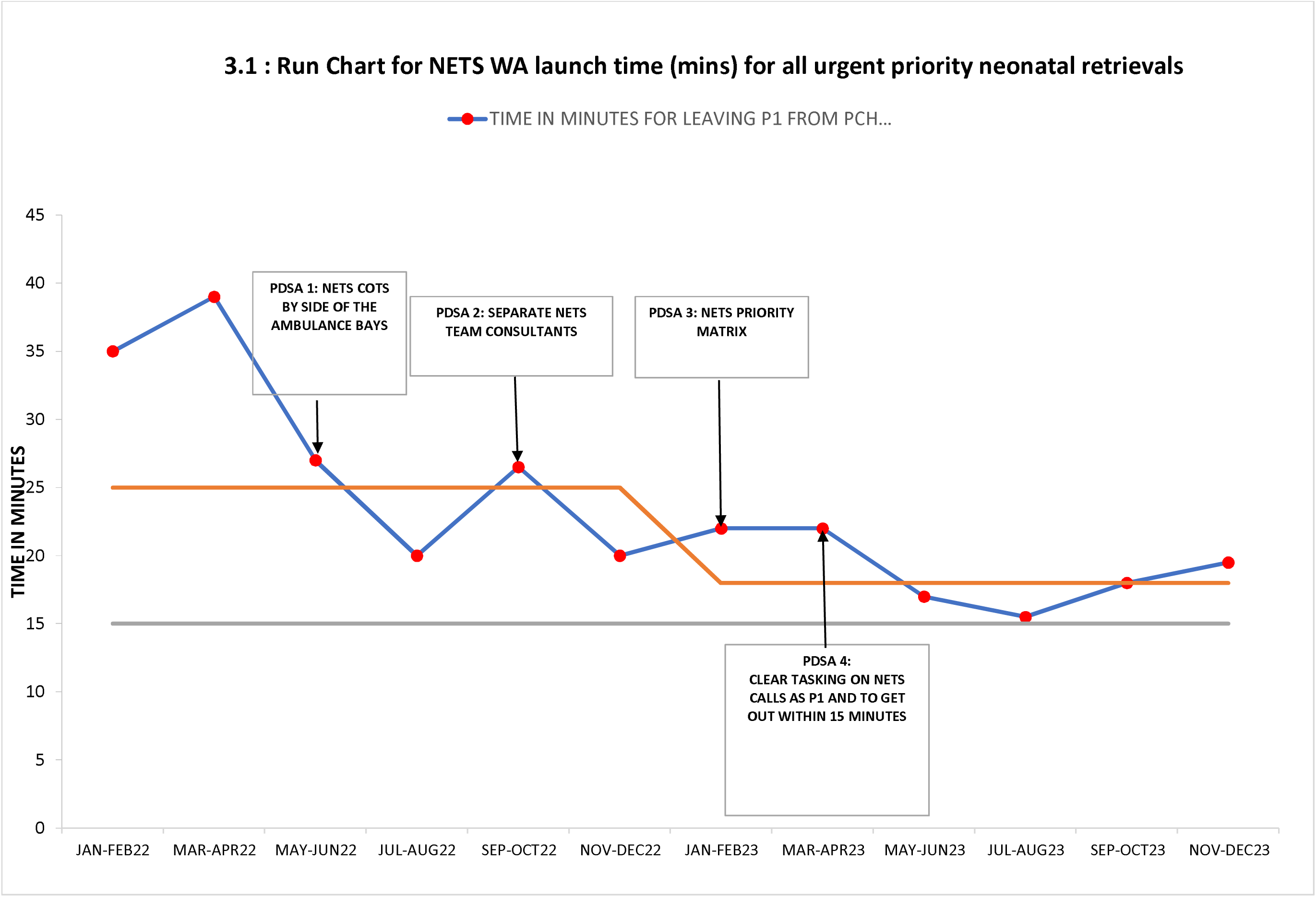
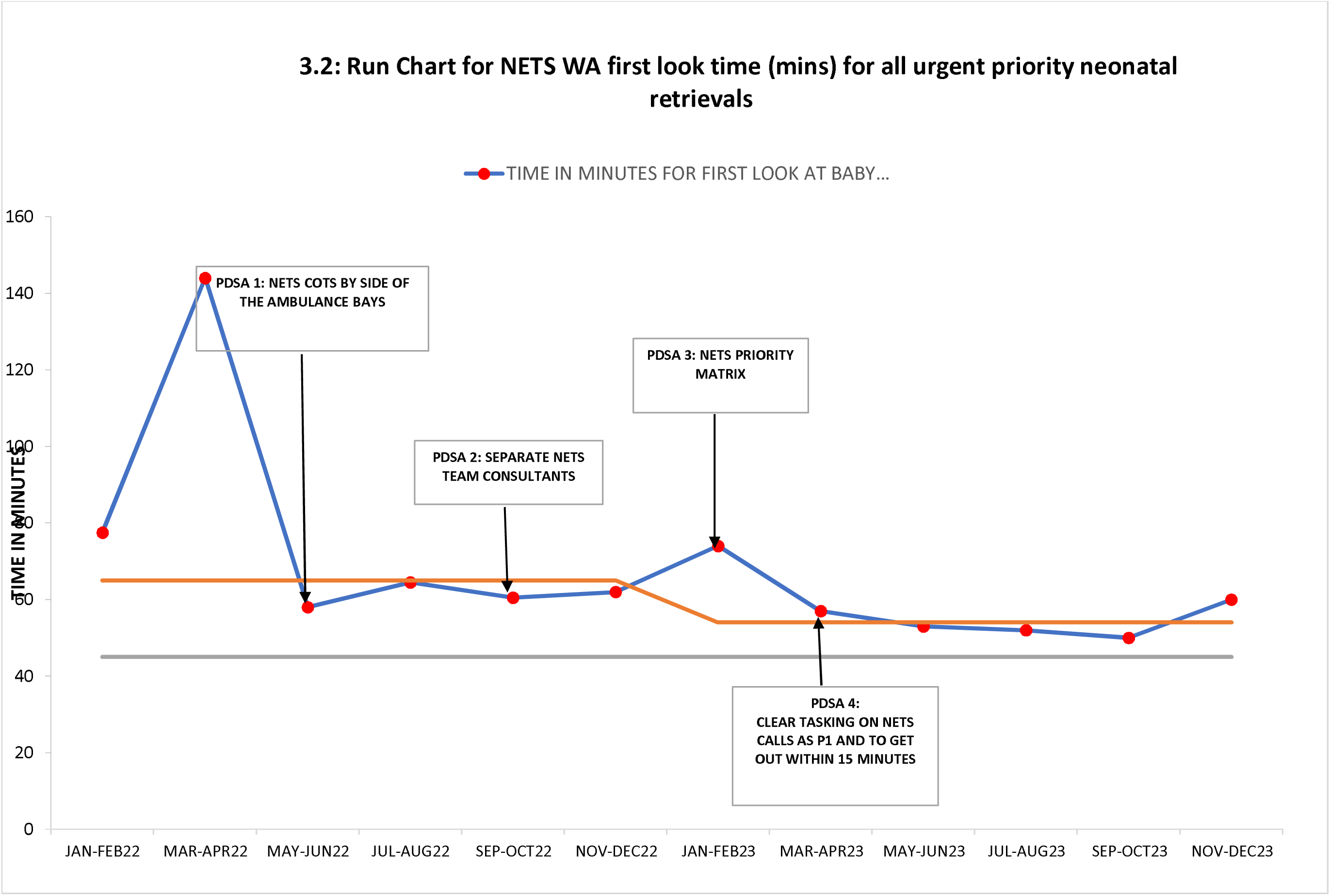
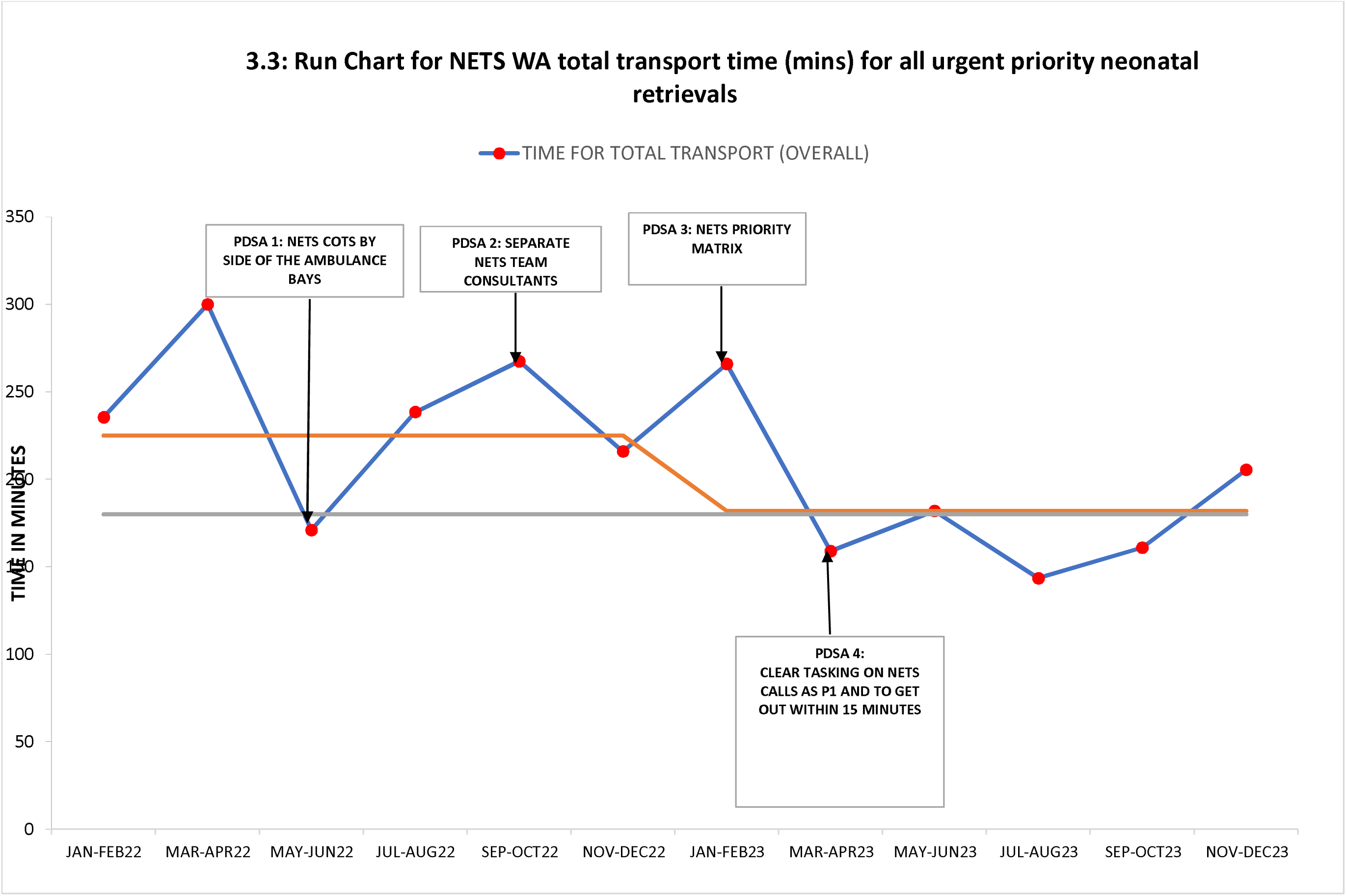
Run-charts

## Results

NETS WA transported 1206 and 1120 retrievals in 2022 and 2023, respectively. The proportion of urgent priority retrievals increased from 110/1206 (9.12%) in 2022 to (172/1120 (15.35%) in 2023 (p<0.0001).

Launch time, first look time and total retrieval time are depicted below in tables 1-3. These describe the metrics for all (table 1), road-only (table 2; metro) and road and air (table 3; regional) urgent priority retrievals respectively. Tables depict data collection for time-centric metrics during baseline (January-May 2022 ie 5 months), during PDSA cycles (June 2022-April 2023 ie 11 months) and post the PDSA cycles (May-December 2023 ie 8 months).

**Table 1.** This table is for all (metro and regional) urgent priority NETS WA retrievals.

|  | January –<br>May 2022<br>(1)<br>n=42 | June 2022 –<br>April 2023<br>(2)<br>n=121 | May –<br>December 2023<br>(3)<br>n=119 | p |
| --- | --- | --- | --- | --- |
| Gestational age at birth<br>(completed weeks) | 38.5<br>(34.7 – 40.2) | 38.2<br>(32.5 - 39.4) | 38.4<br>(35.1 – 40.0) | 0.42 |
| Birth weight<br>(g) | 3095<br>(2275 – 3493) | 3000<br>(1964 – 3500) | 3095<br>(2024 – 3513) | 0.96 |

| Retrieval reason |  |  |  |  |
| --- | --- | --- | --- | --- |
| Non-Surgical | 39 | 99 | 94 | 0.21 |
| Surgical | 3 | 22 | 25 |  |
| Time taken to leave base.<br>(minutes) | 35.5<br>(21.5 – 90.0) | 22.0<br>(13.0 – 45.0) | 17.0<br>(11.0 – 37.0) | **0.0006\*** |
| Time from decision to retrieve to first look.<br>(minutes) | 85.0<br>(54.8 – 269.3) | 62.0<br>(40.0 – 170.5) | 52.5<br>(30.5 – 152.3) | **0.008<sup>§</sup>** |
| Total time of transport<br>(minutes) | 243.5<br>(135.8 – 395.3) | 218.0<br>(136.0 – 385.0) | 182.0<br>(117.0 – 390.0) | 0.33 |

|  | January –<br>May 2022<br>(1)<br>n= 29 | June 2022 –<br>April 2023<br>(2)<br>n=84 | May –<br>December 2023<br>(3)<br>n=91 | p |
| --- | --- | --- | --- | --- |

\* Significant p values in multiple comparisons: 1 vs 2 = 0.02; 1 vs 3 = 0.0004
<sup>§</sup>Significant p values in multiple comparisons: 1 vs 3 = 0.006

**Table 2.**
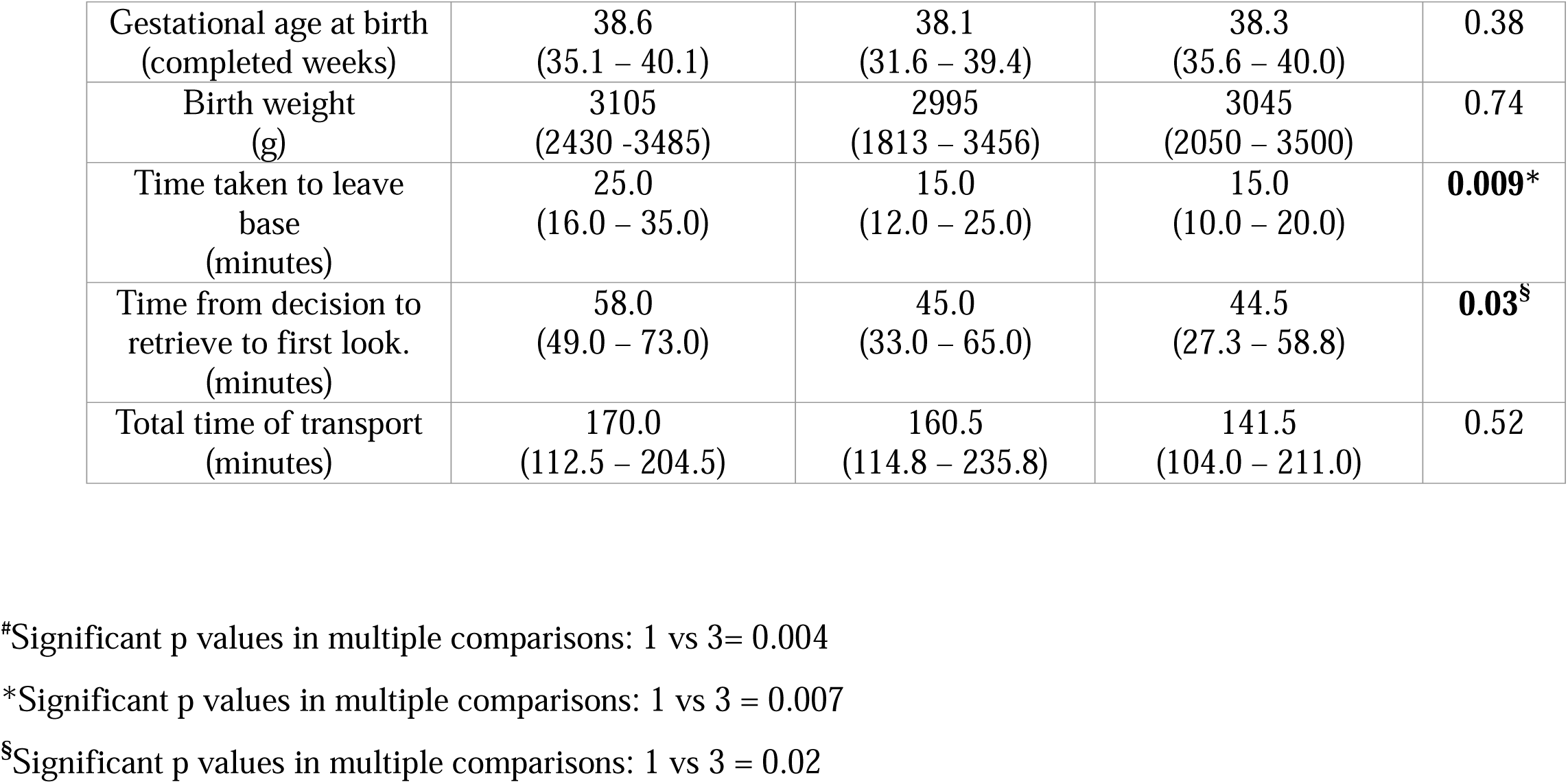
This table is for urgent priority NETS WA retrievals needing only road (metro) transfers. Categorical data is presented as n (%) and compared across all groups by Fisher’s exact test. Continuous data is presented as median (IQR). Groups (1-4) were compared by non-parametric means (Kruskal-Wallis) with Dunn’s correction for multiple comparisons.

|  |  |  |  |  |
| --- | --- | --- | --- | --- |
| Gestational age at birth<br>(completed weeks) | 38.6<br>(35.1 – 40.1) | 38.1<br>(31.6 – 39.4) | 38.3<br>(35.6 – 40.0) | 0.38 |
| Birth weight<br>(g) | 3105<br>(2430 -3485) | 2995<br>(1813 – 3456) | 3045<br>(2050 – 3500) | 0.74 |
| Time taken to leave<br>base<br>(minutes) | 25.0<br>(16.0 – 35.0) | 15.0<br>(12.0 – 25.0) | 15.0<br>(10.0 – 20.0) | **0.009\*** |
| Time from decision to<br>retrieve to first look.<br>(minutes) | 58.0<br>(49.0 – 73.0) | 45.0<br>(33.0 – 65.0) | 44.5<br>(27.3 – 58.8) | **0.03<sup>§</sup>** |
| Total time of transport<br>(minutes) | 170.0<br>(112.5 – 204.5) | 160.5<br>(114.8 – 235.8) | 141.5<br>(104.0 – 211.0) | 0.52 |

#Significant p values in multiple comparisons: 1 vs 3= 0.004
\*Significant p values in multiple comparisons: 1 vs 3 = 0.007
§Significant p values in multiple comparisons: 1 vs 3 = 0.02

**Table 3.**
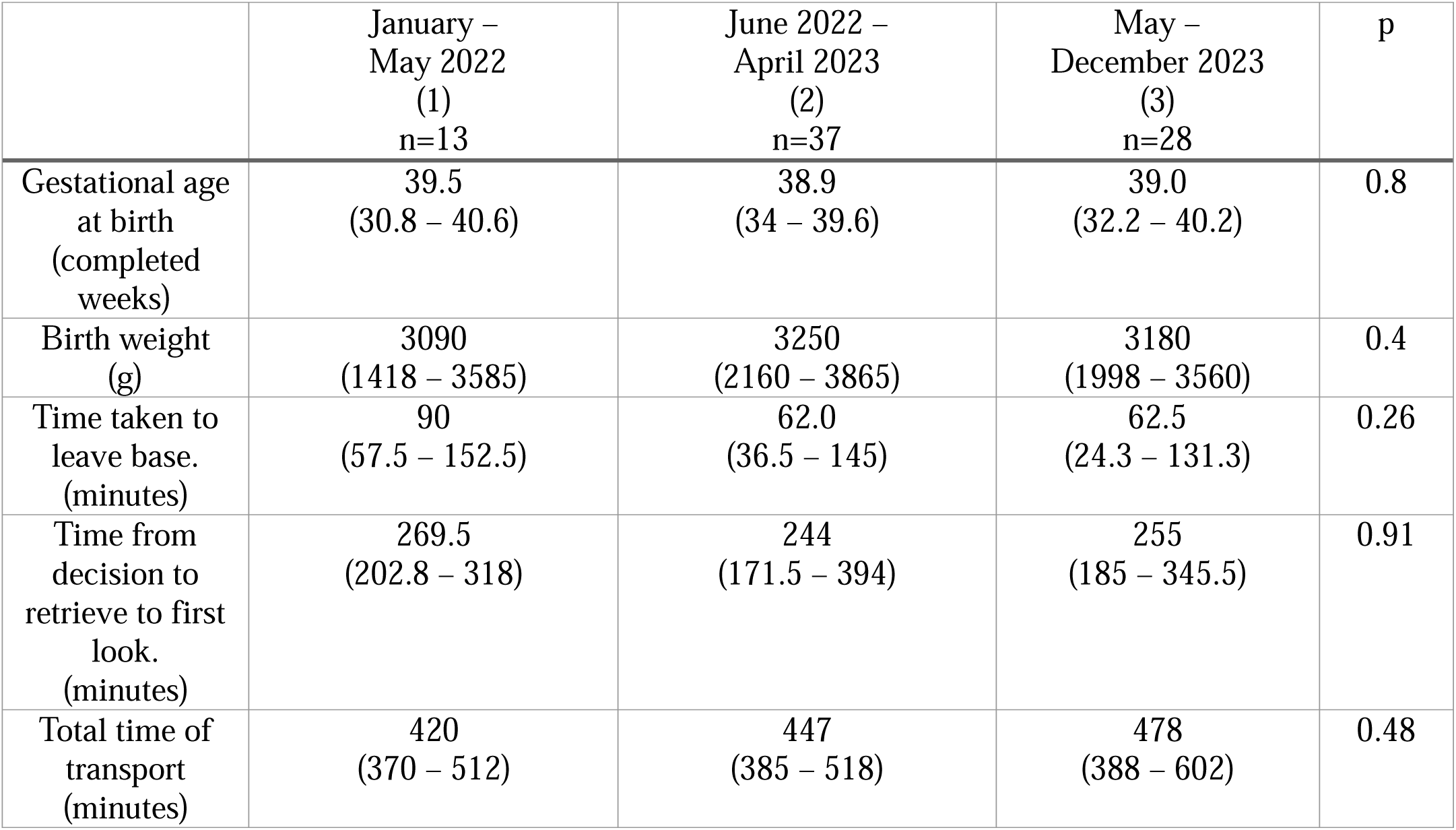
This table is for urgent priority NETS WA retrievals needing air (fixed wing/rotary wing aircraft) + road (regional) transfers. Categorical data is presented as n (%) and compared across all groups by Fisher’s exact test. Continuous data is presented as median (IQR). Groups (1-3) were compared by non-parametric means (Kruskal-Wallis) with Dunn’s correction for multiple comparisons. Significant values are highlighted in bold. Significant comparisons are detailed below the table.

|  | January –<br>May 2022<br>(1)<br>n=13 | June 2022 –<br>April 2023<br>(2)<br>n=37 | May –<br>December 2023<br>(3)<br>n=28 | p |
| --- | --- | --- | --- | --- |
| Gestational age<br>at birth<br>(completed<br>weeks) | 39.5<br>(30.8 – 40.6) | 38.9<br>(34 – 39.6) | 39.0<br>(32.2 – 40.2) | 0.8 |
| Birth weight<br>(g) | 3090<br>(1418 – 3585) | 3250<br>(2160 – 3865) | 3180<br>(1998 – 3560) | 0.4 |
| Time taken to<br>leave base.<br>(minutes) | 90<br>(57.5 – 152.5) | 62.0<br>(36.5 – 145) | 62.5<br>(24.3 – 131.3) | 0.26 |
| Time from<br>decision to<br>retrieve to first<br>look.<br>(minutes) | 269.5<br>(202.8 – 318) | 244<br>(171.5 – 394) | 255<br>(185 – 345.5) | 0.91 |
| Total time of<br>transport<br>(minutes) | 420<br>(370 – 512) | 447<br>(385 – 518) | 478<br>(388 – 602) | 0.48 |

Median (IQR) launch times reduced from 35.5 (21.5-90.0) minutes at baseline to 17.0 (11.0-37.0) (p=0.0006) minutes in the post PDSA cycle period for all urgent priority retrievals. Similarly, median (IQR) first look times reduced from of 85.0 (54.8-269.3) minutes at baseline to 52.5 (30.5-152.3) minutes (p=0.008) in the post PDSA cycle period for all urgent priority retrievals (table 1).

Median launch times for all road only (metro) urgent retrievals reduced to the recommended^7^median (IQR) duration of 15.0 (10.0-20.0) minutes (p=0.009) in the post PDSA cycle period. Median (IQR) first look time reduced from 58.0 (49.0-73.0) minutes at baseline to 44.5 (27.3-58.8) minutes (p=0.03) in the post PDSA cycle period for all road only (metro) urgent retrievals. Total median (IQR) retrieval times reduced from 170.0 (112.5-204.5) minutes to 141.5 (104.0-211.0) minutes (p=0.52) for all road only (metro) urgent retrievals (p=0.52) (table 2).

For air and road (regional) urgent priority retrievals median (IQR) launch times reduced from 90.0 (57.5-152.5) minutes at baseline to 62.5 (24.3-131.3) minutes in the post PDSA cycles period (p=0.26). Median (IQR) first look time reduced from 269.5 (202.8-318) minutes at baseline to 255 (185-345.5) minutes in the post PDSA cycle period (p= 0.91) (table 3).

Categorical data is presented as n (%) and compared across all groups by Fisher’s exact test. Continuous data is presented as median (IQR). Groups (1-3) were compared by non-parametric means (Kruskal-Wallis) with Dunn’s correction for multiple comparisons. Significant values are highlighted in bold. Significant comparisons are detailed below the table.

### Run charts

The Run chart depicts the launch times for all urgent priority retrievals as median (IQR) over two monthly intervals in the study period (figure 3). The run charts include the impact of quicker team launch times on other essential time-centric quality metrics (first look time and total retrieval time) (figure 3).

## Discussion

The present QI study demonstrated the impact of well-planned PDSA cycles by NETS WA team to improvise their time-centric quality metrics on urgent priority neonatal retrievals. Most importantly, these QI measures helped NETS WA team to achieve the desired “SMART goal” of quicker launch times for urgent priority metro retrievals to 15 minutes or less, recommended by the ANZNRN 2022 data dictionary (tables 1-2 figure 3) ^7^. Quicker launch times were the most effective for road only retrievals within the metropolitan area, accounting for majority of the NETS WA retrievals (∼70%) (table 2, figure 3). Quicker launch times had a positive impact on the reduction of “first look time” and “total retrieval time”. Quicker first look time translated into NETS WA teams having earlier access to sick infants for provision of urgent time-critical care, lacking in those referring units. Reduced total retrieval time meant these sick infants received tertiary level care earlier, potentially favorably altering their clinical course (tables 1-3, figure 3).

Globally, some neonatal transport networks have recently developed time-centric quality metrics for benchmarking. One of the most important quality metrics, unanimously endorsed is the team activation time referred to as mobilization or launch time ^5–8^. Quicker launch times enable retrieval teams with rapid access to sick infants needing higher-level of care. Targeting quicker launch times is more practical than stabilization times in neonatal retrievals. This is due to the unique newborn physiology necessitating prolonged stabilization^9, 18^. Further, shorter stabilization times in adult and pediatric retrievals fail to yield similar neonatal outcomes ^8, 19–21^. Hence, QI studies to achieve quicker launch times would be more beneficial in time-critical neonatal retrievals. However, it is challenging to perform QI studies in neonatal retrieval settings because of its dynamic nature.

Some recent effective QI measures in neonatal retrieval settings include reducing brain injury in extreme preterm infants and thermoregulation in outborn infants^22, 23^. There is a dearth of QI studies targeting launch times in urgent priority neonatal retrievals. Rajapreyar et al. reported an improvement in the mobilization times of their retrieval teams to less than 25 minutes. They did focused interventions such as recognizing the team with the quickest mobilization times and documenting reasons for delay ^10^. The study involved road and rotary wing retrievals but no fixed wing aircraft retrievals. The study included both pediatric and neonatal retrievals with a mix of respiratory therapist or physician and a registered nurse ^10^. In contrast, NETS WA have a more uniform approach of dispatching a senior neonatal registrar (advanced neonatal trainee) and/or consultant with a senior nurse for all urgent retrievals. Further, NETS WA transfers have a mix of road, fixed and rotary wing aircraft retrievals. Another QI study by Arcinue et al targeted the mobilization time of their retrieval teams to less than 30 minutes^4^. Their PDSA cycles involved better communication skills, open critical bed space for transfers, faster patient room turnovers, more teams and involving the accepting neonatologist with the referring physician and transport physician on the initial call. The study showed success of these QI measures with the target mobilization time achieved in 82% of retrievals compared to 27% at baseline^4^. Kenningham et al. included 2 PDSA cycles with phase 1 involving separate dedicated retrieval team and phase 2 with process mapping. The study reported a 20% reduction of their mobilization time from baseline for road only transfers^24^. The present study differs from the above two studies as NETS WA focused on the “urgent” priority retrievals needing expedited launch times (<15 minutes) for the most time-critical conditions via road and/or air, over the largest retrieval area in the world. Further, the present study showed the impact of quicker launch times with an improvement in other essential time-centric quality metrics such as “first look time” and “total retrieval time”.

The present study has several strengths. Itis the first conducted in an Australasian neonatal retrieval setting and provides baseline data for time-centric quality metrics (ANZNRN 2022)^7^. This is helpful for other national and international neonatal retrieval teams for benchmarking. The SMART goal of targeting launch times (less than 15 minutes) was achieved within a reasonable time frame of two years. The impact of quicker launch times on improving other important time-centric quality metrics (first look time and total retrieval time) adds essential clinical relevance for urgent priority neonatal retrievals. Results were statistically significant and sustained over a long period after the last PDSA cycle. This adds credibility to the impact of the introduced QI measures. Study QI measures are generalizable and can be easily adapted by other neonatal retrieval teams. The other strengths include standardized electronic data collection, regular FGD with education sessions and strict adherence to the SQUIRE 2.0 guidelines ^15^.

Limitations of this reported process was the lesser impact of the QI measures on non-metro retrievals. These retrievals are dependent on multiple non-modifiable factors such as availability of resources (aircraft, crew), aeromedical (pilot hours, civil aviation regulations) and weather constraints. These metrics are less amenable for modifications by any QI measures. Another limitation was lack of assessment of impact of quicker launch times on scene/stabilization time. Reducing launch times is likely more beneficial than stabilization times for neonatal retrievals attributed to their unique physiology^9, 19, 20^. The lack of investigations around the astronomical data points in the run charts is another study limitation.

The future implications include regular PDSA cycles to ascertain the sustenance of these improved time metrics with investigations for the astronomical data points. There is scope for inter-agency collaboration to improvise time metrics for non-metro retrievals. Studies could target the impact of improved time metrics in terms of clinical outcomes against a historical cohort. Further, it would be useful to compare these quality metrics with other ANZNRN and international neonatal retrieval teams for benchmarking purposes.

To conclude, well-designed QI measures can quicken the benchmarking time-centric quality metrics of neonatal transport teams on urgent priority retrievals.

## Data Availability

All data produced in the present study are available upon reasonable request to the authors

## Bibliography

1. Lupton BA and Pendray MR. Regionalized neonatal emergency transport. Semin Neonatol 2004; 9: 125–133. DOI: 10.1016/j.siny.2003.08.007.

2. Leslie A and Fenton A. Categorising neonatal transports. Arch Dis Child Fetal Neonatal Ed 2012; 97: F77. 20111012. DOI: 10.1136/archdischild-2011-300509.

3. Rowley R PM, Thwaites R. What Proportion of Transfers is Time Critical and are we Mobilising Quickly Enough. Neonatal society 2013 summer meeting. Edinburgh 2013.

4. Arcinue R, Chapman RL, Van-Allen K, et al. Improving Transport Team Mobilization for Emergent Neonatal Transfers: A Transport-Access Center-Neonatology (TAN) Quality Improvement Initiative. Pediatrics 2021; 147: 1025–1025. DOI: 10.1542/peds.147.3MA10.1025a.

5. Bigham MT and Schwartz HP. Quality metrics in neonatal and pediatric critical care transport: a consensus statement. Pediatr Crit Care Med 2013; 14: 518–524. DOI: 10.1097/PCC.0b013e31828a7fc1.

6. Schwartz HP, Bigham MT, Schoettker PJ, et al. Quality Metrics in Neonatal and Pediatric Critical Care Transport: A National Delphi Project. Pediatr Crit Care Med 2015; 16: 711–717. DOI: 10.1097/pcc.0000000000000477.

7. Network AaNZN. Australian and New Zealand Neonatal Retrieval Minimum Dataset and Data Dictionary: Pilot. Australian and New Zealand Neonatal Retrieval Minimum Dataset and Data Dictionary: Pilot. 1 ed. 2022, p. 88.

8. Lee KS. Neonatal transport metrics and quality improvement in a regional transport service. Transl Pediatr 2019; 8: 233–245. DOI: 10.21037/tp.2019.07.04.

9. Whitfield JM and Buser MK. Transport stabilization times for neonatal and pediatric patients prior to interfacility transfer. Pediatr Emerg Care 1993; 9: 69–71. DOI: 10.1097/00006565-199304000-00002.

10. Rajapreyar P, Badertscher N, Willie C, et al. Improving Mobilization Times of a Specialized Neonatal and Pediatric Critical Care Transport Team. Air Medical Journal 2022; 41: 315–319.

11. Australian Bureau of Statistics. 1306.5 - Western Australia at a Glance, 2024.

12. Thompson K, Gardiner J and Resnick S. Outcome of outborn infants at the borderline of viability in Western Australia: A retrospective cohort study. J Paediatr Child Health 2016; 52: 728–733. 20160505. DOI: 10.1111/jpc.13187.

13. Davis JW, Seeber CE, Nathan EA, et al. Outcomes to 5 years of outborn versus inborn infants <32 weeks in Western Australia: a cohort study of infants born between 2005 and 2018. Arch Dis Child Fetal Neonatal Ed 2023; 108: 499–504. 20230217. DOI: 10.1136/archdischild-2022-324749.

14. Gardiner J, McDonald K, Blacker J, et al. Unintended events in long-distance neonatal interhospital transport in Western Australia: a comparison of neonatal specialist and non-neonatal specialist transport teams. The Journal of Pediatrics: Clinical Practice 2024; 11: 200102.

15. Ogrinc G, Davies L, Goodman D, et al. SQUIRE 2.0 (Standards for QUality Improvement Reporting Excellence): revised publication guidelines from a detailed consensus process. BMJ Qual Saf 2016; 25: 986–992. 20150914. DOI: 10.1136/bmjqs-2015-004411.

16. Pronovost P, Needham D, Berenholtz S, et al. An intervention to decrease catheter-related bloodstream infections in the ICU. N Engl J Med 2006; 355: 2725–2732. DOI: 10.1056/NEJMoa061115.

17. Taylor MJ, McNicholas C, Nicolay C, et al. Systematic review of the application of the plan-do-study-act method to improve quality in healthcare. BMJ Qual Saf 2014; 23: 290–298. 20130911. DOI: 10.1136/bmjqs-2013-001862.

18. Chakkarapani AA, Whyte HE, Massé E, et al. Procedural Interventions and Stabilization Times During Interfacility Neonatal Transport. Air Med J 2020; 39: 276–282. 20200507. DOI: 10.1016/j.amj.2020.04.007.

19. McPherson ML and Graf JM. Speed isn’t everything in pediatric medical transport. Pediatrics 2009; 124: 381–383. DOI: 10.1542/peds.2008-3596.

20. Orr RA, Felmet KA, Han Y, et al. Pediatric specialized transport teams are associated with improved outcomes. Pediatrics 2009; 124: 40–48. DOI: 10.1542/peds.2008-0515.

21. Boden WE, Eagle K and Granger CB. Reperfusion strategies in acute ST-segment elevation myocardial infarction: a comprehensive review of contemporary management options. J Am Coll Cardiol 2007; 50: 917–929. 20070821. DOI: 10.1016/j.jacc.2007.04.084.

22. Mohammad K, Momin S, Murthy P, et al. Impact of quality improvement outreach education on the incidence of acute brain injury in transported neonates born premature. J Perinatol 2022; 42: 1368–1373. 20220504. DOI: 10.1038/s41372-022-01409-2.

23. Redpath S, Moore H, Sucha E, et al. Therapeutic Hypothermia on Transport: The Quest for Efficiency: Results of a Quality Improvement Project. Pediatr Qual Saf 2022; 7: e556. 20220614. DOI: 10.1097/pq9.0000000000000556.

24. Kenningham K, Billimoria Z, Baker C, et al. Effect of Neonatal Transport Team Structure And Activation Process Revisions On Team Mobilization Time. Pediatrics 2021; 147: 1015–1016. DOI: 10.1542/peds.147.3MA10.1015.

