## appendix for "Quicker team launch times for urgent priority neonatal retrievals: A Quality Improvement Initiative study"

Appendix 1: Standard NETS WA Process for retrievals:

1. Phone call received to statewide free-call 1300 number with automated recording; answered by NETS WA nurse or senior registrar on call
2. Isobar handover taken including recording of demographics of patient/s
3. Call patched to NETS WA consultant on call with referring site and senior registrar. Collaborative planning, advising, problem solving occurs over 3-way chat.
4. Designation of plan with cessation of call and NETS WA communication to 3rd parties i.e. SJA and/or RFDS for metropolitan and non-metropolitan retrievals.
5. Departure of NETS WA team from base.
6. Arrival at the referring hospital and first look at the baby
7. Stabilization of the infant and call-back to NETS WA base to communicate the status
8. Transport to the sole tertiary referral centres in Perth, WA.

Appendix 2: Transport variables for priority 1 (P1) retrievals

1. Date of birth
2. Birth gestation (weeks)
3. Birth weight (grams)
4. Reason for retrieval: Medical or surgical
5. Area of retrieval: Metropolitan or non-metropolitan area
6. Mode of transport: road only or road + fixed wing aircraft or road + rotary wing aircraft
7. Ambulance services: NETS WA or SJA
8. Date and time of decision to retrieve: Task time
9. Date and time leaving the ambulance bay
10. Total time taken to leave ambulance bay from time of tasking: Launch time (minutes)
11. Date and time of arrival of NETS WA team for first look at the baby
12. Total time taken from task time till first look of the baby: First look time (minutes)
13. Total time taken from task time to come back at base: Total retrieval time (minutes)

Appendix 3: Neonatal conditions for routinely activating P1 tasking of NETS WA teams:

1. Prematurity less than 30+^0^ weeks gestational age at birth
2. Severe respiratory distress needing mechanical ventilation with oxygen requirement >60% +/- need for inhaled nitric oxide.
3. Shock or acute cardiovascular collapse +/- duct dependent cardiac conditions needing additional medications (inotropes, prostaglandins)
4. Moderate to severe Hypoxic Ischemic Encephalopathy needing therapeutic hypothermia.
5. Seizures needing more than 2 anticonvulsant medications.
6. Severe Jaundice needing exchange transfusion.
7. Urgent surgical conditions: intestinal perforation, bilious vomiting to rule out intestinal malrotation, testicular torsion.
8. Suspected inborn errors of metabolism (eg ammonia> 100 mg/l)
